# Improving prediction models of amyotrophic lateral sclerosis (ALS) using polygenic, pre-existing conditions, and survey-based risk scores in the UK Biobank

**DOI:** 10.1101/2024.03.28.24305037

**Authors:** Weijia Jin, Jonathan Boss, Kelly M. Bakulski, Stephen A. Goutman, Eva L. Feldman, Lars G. Fritsche, Bhramar Mukherjee

**Author notes:** These authors contributed equally to this work (Lars G. Fritsche, Bhramar Mukherjee).

## Abstract

**Background and Objectives:** Amyotrophic lateral sclerosis (ALS) causes profound impairments in neurological function and a cure for this devastating disease remains elusive. Early detection and risk stratification are crucial for timely intervention and improving patient outcomes. This study aimed to identify predisposing genetic, phenotypic, and exposure-related factors for Amyotrophic lateral sclerosis using multi-modal data and assess their joint predictive potential.

**Methods:** Utilizing data from the UK Biobank, we analyzed an unrelated set of 292 ALS cases and 408,831 controls of European descent. Two polygenic risk scores (PRS) are constructed: “GWAS Hits PRS” and “PRS-CS,” reflecting oligogenic and polygenic ALS risk profiles, respectively. Time-restricted phenome-wide association studies (PheWAS) were performed to identify pre-existing conditions increasing ALS risk, integrated into phenotypic risk scores (PheRS). A poly-exposure score (“PXS”) captures the influence of environmental exposures measured through survey questionnaires. We evaluate the performance of these scores for predicting ALS incidence and stratifying risk, adjusting for baseline demographic covariates.

**Results:** Both PRSs modestly predicted ALS diagnosis, but with increased predictive power when combined (covariate-adjusted receiver operating characteristic [AAUC] = 0.584 [0.525, 0.639]). PheRS incorporated diagnoses 1 year before ALS onset (PheRS1) modestly discriminated cases from controls (AAUC = 0.515 [0.472, 0.564]). The “PXS” did not significantly predict ALS. However, a model incorporating PRSs and PheRS1 improved prediction of ALS (AAUC = 0.604 [0.547, 0.667]), outperforming a model combining all risk scores. This combined risk score identified the top 10% of risk score distribution with a 4-fold higher ALS risk (95% CI: [2.04, 7.73]) versus those in the 40%-60% range.

**Discussions:** By leveraging UK Biobank data, our study uncovers predisposing ALS factors, highlighting the improved effectiveness of multi-factorial prediction models to identify individuals at highest risk for ALS.

## Introduction

Amyotrophic lateral sclerosis (ALS), also known as Lou Gehrig’s disease, is a form of Motor Neurone Disease (MND), affecting the nerve cells (neurons) responsible for controlling voluntary muscle movement [1-3]. ALS is a rare neurodegenerative disease with an incidence of approximately 2 per 100,000 person-years, and a prevalence of 6–9 per 100,000 persons globally [1-3]. ALS remains incurable till today and patients only survive 2 to 5 years post-diagnosis [1-4]. Current management is centered on medications that only moderately delay disease progression, and on supportive equipment that improve the quality of life for patients. Given these considerations, risk stratification and early ALS detection can expedite treatment initiation to slow progression and relieve disease burden [2].

Multiple demographic, genetic, environmental, and phenotype factors correlate with ALS [5, 6]. For instance, older age, male sex, and a family history of ALS raise the risk for ALS [2, 7, 8]. Studies have also identified genetic risk, with over 40 ALS genes, including C*9orf72*, *SOD1*, *FUS*, and *TARDBP* [9-11], as well as environmental risks from exposures (e.g., to metals, pesticides, intense physical activity [5, 6, 12]). Despite these findings and the resulting advances in ALS correlation and prediction models [13-15], a noticeable gap persists in the holistic ALS risk stratification literature due to a lack of approaches incorporating integrated sets of multi-domain risk factors into ALS prediction models [16]. This highlights the need for more comprehensive and integrated approaches to enhance the accuracy and reliability of ALS risk prediction.

This study aims to bridge this gap by investigating a comprehensive set of ALS risk predictors by integrating multi-modal data from a cohort of over 500,000 people recruited between 2006 and 2010 from the UK Biobank, a large-scale biological database [17]. The ALS analytic cohort included 292 participants with a reported ALS diagnosis in their electronic health records (EHRs) or self-reported survey data.

We explored three types of ALS risk scores: Polygenic Risk Scores (PRS [18]), Poly Exposure Risk Scores (PXS [19]), and Phenotype Risk Scores (PheRS [20]), derived from genome-wide association studies (GWAS), self-reported survey data, and phenome-wide association studies (PheWAS), using the UK Biobank. The PRS reflects germline genetics, PXS health-related exposure, and PheRS historical clinical data. In our analyses, we adjusted for factors such as sex, age, genotyping, and PCA variables. Age and sex, are established risk factors for ALS [21, 22] and we evaluated these predictors separately and jointly to predict and stratify ALS risk.

Our study seeks to refine ALS risk prediction using a comprehensive data platform like the large-scale UK Biobank, potentially contributing to early detection and prevention strategies for high-risk individuals, thereby advancing our understanding and management of ALS. The relative predictive importance of each data domain can also guide priorities in future data collection and harmonization efforts.

## Methods

### Subjects

This study employed data from the UK Biobank (UKB), a population-based cohort collected from multiple sites across the United Kingdom and includes over 500,000 participants aged between 40 and 69 years when recruited in 2006–2010 [17]. For this study, various types of UK Biobank data were utilized, including questionnaire data, electronic health record (EHR) data, and genotype and genotyped-derived data.

### Genotype data

In the data cleaning process, we excluded 2,331 samples flagged by the UK Biobank quality control (QC) documentation [23], leaving 485,416 individuals post-sample QC filtering. We used the UK BioBank Imputed Dataset (v3, see **Web Resources**) and limited analyses to variants with imputation information score >= 0.3 and minor allele frequency (MAF) >= 0.01%.

The online augmentation, decomposition, and Procrustes (OADP) method was applied to the genotype data of UK Biobank samples with 2,492 samples from the 1000 Genomes Project data as the reference (FRAPOSA; see **Web Resources**) to infer ancestry, and super populations membership (AFR: African, AMR: Ad Mixed American, EAS: East Asian, EUR: European, SAS: South Asian ancestry) [24]. For this study, genetic principal components were calculated within the EUR analytic sample.

### Outcome and covariate data

427 cases of MND were identified utilizing the following: ICD9 (fields: 40013, 41203, and 41205; code 335.2), ICD10 (fields: 40001, 40002, 40006, 41201, 41202, and 41204; code G12.2) and self-reported “motor neurone disease” (field: 20002; code 1259) (see **Web Resources**). The covariates utilized in our analyses include age (see below), genotyping array (UK Biobank Lung Exome Variant Evaluation or [UK BiLEVE] Axiom array [25]), the first four principal components from the genotype data (PC1 – PC4; field: 22009), and genetically inferred sex (field: 22001). Despite the synonymous use of ‘motor neurone disease’ (MND) and ALS in the UK[26], we refer to these cases as ‘ALS’ for clarity, although not all MND cases are classified as ALS. A flowchart of the process of case-control selection is captured in **Figure 1 and Figure S1** and described below: Age of ALS onset was defined as the earliest age at first recorded MND ICD diagnosis or the self-reported age of onset. We excluded cases that failed genotype QC (n = 10) and those lacking diagnosis-based or self-reported age of onset (n = 33). Further exclusions included ALS age of onset under 18 (n = 4), onset before the last assessment (n = 82), and inferred non-EUR ancestry (n = 6), leaving 292 of inferred European ancestry (EUR). All cases were unrelated and had exposure data available.

**Figure 1.**
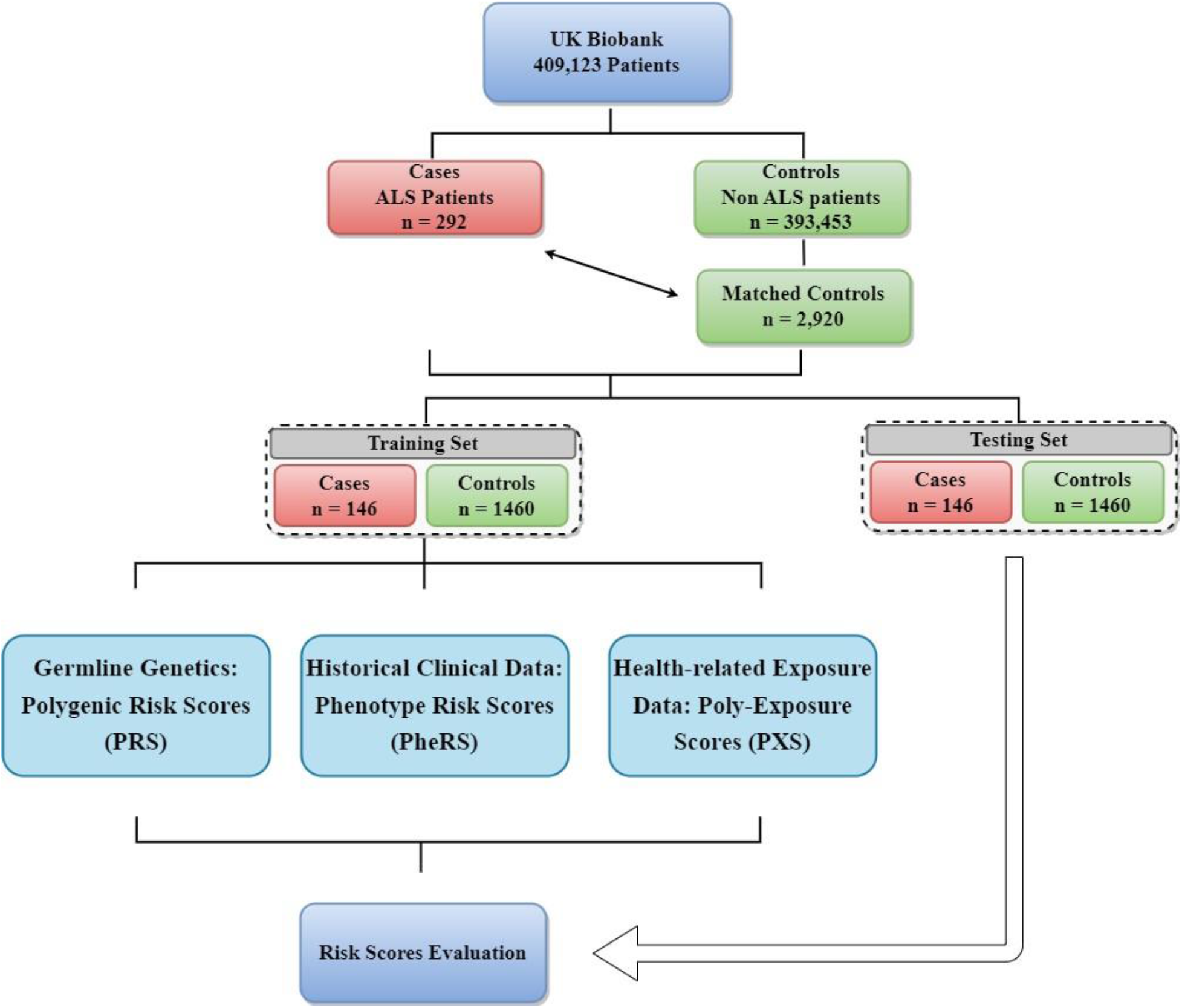
Overview flowchart illustrating analysis setup. Details about case and control definitions as well as sample filtering can be found in the Method section and **Figure S2**.

484,999 samples that passed genotype QC and had no ALS diagnosis were used for control selection. Further, we excluded individuals diagnosed with any hereditary/degenerative nervous conditions (phecodes 330–337.9) or other diseases of the central nervous system (phecodes 341– 349.9) and individuals who self-reported having myasthenia gravis (code 1260), multiple sclerosis (code 1261), Parkinson’s disease (code 1262), or dementia/Alzheimer’s/cognitive impairment (Field 20002: code 1263). This combined step led to the exclusion of 20,058 samples.

Subsequently, to ensure uniformity in genetic background, individuals not of genetically inferred European ancestry were removed, resulting in the exclusion of another 25,231 samples. We then removed 20 samples with missing exposure data. To minimize genetic heterogeneity or cryptic relatedness, individuals with a kinship coefficient of > 0.0884 were removed, leading to the exclusion of 30,859 samples [27]. This process entailed selecting a maximal set of unrelated controls unrelated to the previously selected cases [28].

Propensity score matching using R package MatchIt was then performed based on age, the first four PCs from the genotype data (PC1-PC4), a caliper of 0.25, and exact matching on sex, with a matching ratio allowing a maximum of 10 controls per case, leading to a set of 2920 matched controls. Finally, the 292 cases were randomly split into two equally sized subsets of 146 to obtain a training and testing cohort; matched controls were retained.

### PRS construction

A PRS aggregates information across a defined set of genetic loci, incorporating each locus’s association with the target trait. The PRS for person *j* takes the form 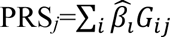 where *i* indexes the included loci for that trait, weight 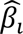 is the estimated log odds ratios retrieved from the external GWAS summary statistics for locus *i,* and *G*_*ij*_ is a continuous version of the measured dosage data for the risk allele on locus *i* in subject *j*.

Summary statistics of a large European ancestry-based GWAS on ALS [29] (also see **Web Resources**) were used to generate two PRS. The first PRS construction method involved linkage disequilibrium (LD) clumping of variants with p-values below 5x10^-8^ using 10,000 randomly selected EUR samples and a pairwise correlation cut-off at r2 < 0.1 within 1 Mb window. Using the remaining variants (“independent GWAS Hits”), the weighted PRS was computed (see above) denoted as “GWAS Hits PRS”. For the second PRS construction method, the software package “PRS-CS” [30] was used to define a PRS based on continuous shrinkage (CS) priors. We applied a MAF filter of 1 % and only included autosomal variants that overlap between summary statistics, LD reference panel, and target panel. For comparison, a recently published ALS PRS based on the same underlying EUR-based GWAS on ALS and included 275 uncorrelated single-nucleotide polymorphisms (SNPs) based on an LD clumping and P-value thresholding (C+T) approach, which excludes SNPs from the region around C9orf72[15], was also evaluated, referred to as PRS-275-non-C9. A full list of weights can be downloaded from our website (see **Web Resources**).

The dosage-based value of each PRS was calculated for each UK Biobank individual, utilizing the R package “Rprs” (see Web Resources) and the weights from the three PRS methods.

### Phenotype Risk Score (PheRS)

#### Time-restricted phenome generation

To generate a phecode-based phenome, all available ICD9 and ICD10 data was first extracted for cases and controls. To balance the need for uncovering early signs of ALS and ALS complications, as well as potential sample size reduction due to shorter detection windows, various time thresholds were set to include pre-existing diagnoses recorded more than 0.5, 1, 2, 3, 4, and 5 years before the key date. The key date for cases was the date of the ALS diagnosis, and for controls, the last assessment date or last ICD diagnosis, depending on which occurred first. The extracted ICD codes were aggregated to generate up to 1,814 phecodes using the PheWAS R package [31].

#### Phenome-wide association studies

To identify phenotypes associated with ALS, Firth bias-corrected logistic regression was conducted for each phenotype of the corresponding time-restricted phenomes by fitting the following model.

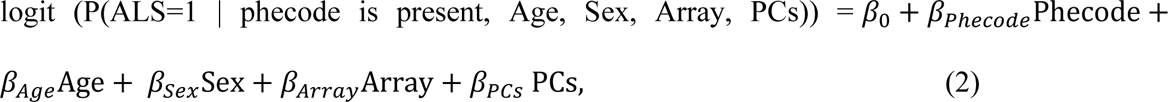

where the PCs represent the first four PCs and “Array” represents the genotyping array.

#### PheRS Construction

To construct a PheRS based on preexisting conditions, we selected the top 50 most significant associations of the time-restricted PheWAS (0.5, 1, 2, 3, 4, or 5 years prior to an ALS diagnosis). We then implemented forward selection using Firth-corrected logistic regression to identify independent phenotypes and multivariable phenotype-adjusted beta estimates [20, 32, 33]. The corresponding PheRS was calculated as the sum of the effect size weighted presence of the corresponding independent set of phenotypes.

The PheRSs for the six time thresholds are referred to as PheRS0.5, PheRS1, PheRS2, PheRS3, PheRS4, and PheRS5, respectively.

### Poly Exposure Score (PXS)

We employed a selection process to identify 111 exposure variables, categorized as indicators of physiological state, environmental exposure, and self-reported behaviors, following He et al.’s approach [34]. Post-processing, we derived 24 continuous, 9 binary, 59 ordered categorical, and 19 unordered categorical variables.

We removed 16 variables demonstrating high levels of missingness (>=20%), perfect correlation with another variable, or deemed non-informative. To address non-random missingness within the dataset (**Supplementary Table S1**), we applied clustered imputation through classification and regression trees (m = 50) via the R package “mice” [35]. To alleviate the impact of multicollinearity and streamline the predictor space, we conducted stepwise variance inflation factor (VIF) filtering, excluding 13 variables consistently demonstrating VIF > 5 (**Supplementary Table S2**), resulting in a total of 82 environmental exposure variables.

The remaining 82 variables were used to construct a ridge-regression-based PXS, adjusting for age, sex, and the first four PCs. Model optimization, guided by cross-validation, selected the optimal lambda for exposure weights. These weights were applied to calculate a weighted PXS for each imputed testing dataset. The final PXS was computed as the mean of the 50 PXS values.

### Statistical analysis

#### Evaluation and identification of important predictive scores

To evaluate each of the generated predictors (PRSs, PXSs, or PheRSs), the following Firth bias-corrected logistic regression model was fit for each PRS on the training data adjusting for covariates age, genotyping array, and the first four PCs:

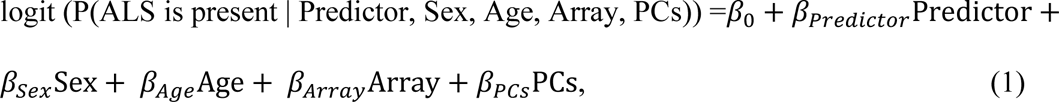

where the PCs represent the first four PCs and “Array” represents the genotyping array.

To evaluate models with multiple predictors (e.g., GWAS hits PRS + PRS-CS predictor), we employed logistic regression on the training set to derive weights for each predictor. These weights were then used to create a weighted combined score applied to the testing data.

For each predictor, the following performance measures were assessed relative to the observed binary ALS status, adjusting for covariates such as age, sex, array, and the first four PCs: overall association and the ability to discriminate cases from controls, as measured by the area under the covariate-adjusted receiver operating characteristic (AROC) curve (denoted AAUC) using R package “ROCnReg” [36]. To compare association effect sizes corresponding to the various predictors, each continuous predictor was centered to its mean and scaled to have a standard deviation of 1.

Finally, a risk stratification analysis was conducted using an aggregate score derived from a select set of predictors. Within the test dataset, the control group’s aggregate score distribution was partitioned into deciles to establish our risk categories. The ALS cases were then allocated into these deciles. To demonstrate the variation in disease risk across deciles, odds ratios (ORs) and their corresponding 95% confidence intervals (CIs) were calculated by fitting logistic regression models, adjusting for covariates, and employing the middle deciles (40-60%) as the reference group.

## Results

### Cohort Characteristics

Among 409,123 participants of the analytical data set of unrelated individuals of European ancestry, 292 (0.07%) had an ALS diagnosis after their last assessment, while 408,831 were considered controls (**Figure S1**). We observed that females were underrepresented among ALS cases (43.5%) versus controls (54.2%). We also noted differences in the age distributions of male and female ALS patients at their first diagnosis (**Figure S2**). Furthermore, we observed that ALS cases were, on average, older (mean age 66.02 years) compared to controls (mean age 61.24 years, **Table 1**). To control for these observed differences in covariates between cases and controls, we performed matching based on age, sex, and the four genotyping-related PCs. This matching approach led to a more balanced distribution of covariates between ALS cases and matched controls (**Table 1**).

**Table 1.** Characteristics of Analytical Sample from UK Biobank. This table presents the features of the analytical sample, specifically 292 ALS cases with onset after assessment and 408831 unmatched and 2920 matched controls. The sample, restricted to genetically inferred European ancestry and unrelated individuals, was selected to minimize genetic heterogeneity and cryptic relatedness. Case-control matching utilized nearest neighbor matching considering age, first four principal components from genotype data (PC1-4), a 0.25 caliper, and exact sex matching, permitting up to 10 matched controls per case. Array type wasn’t a matching criterion but was adjusted for in subsequent analyses.

|  | **ALS Cases** | **Unmatched**<br>**Controls** | **Matched**<br>**Controls** |
| --- | --- | --- | --- |
| n | 292 | 408,831 | 2,920 |
| Age in years, mean (SD) | 66.02 (6.68) | 61.11 (9.07) | 65.99 (6.63) |
| Female (%) | 127 (43.5) | 221649 (54.2) | 1270 (43.5) |
| UK BiLEVE Axiom array (%) | 35 (12.0) | 43956 (10.8) | 335 (11.5) |
| PC1, mean (SD) | -70.07 (1.77) | -69.96 (2.13) | -70.09 (1.67) |
| PC2, mean (SD) | 88.71 (2.01) | 88.75 (2.51) | 88.75 (1.88) |
| PC3, mean (SD) | -18.79 (2.25) | -18.84 (2.44) | -18.82 (2.14) |
| PC4, mean (SD) | 23.17 (1.52) | 23.17 (1.57) | 23.19 (1.47) |

### Germline Genetics: Polygenic Risk Scores (PRS)

Shifting our attention to the genetic factors associated with ALS, we constructed two PRS: “GWAS hits PRS” and “PRS-CS” to investigate the oligogenic and polygenic associations with ALS, respectively. In the test set consisting of 146 ALS cases and 1460 controls, both “GWAS hits PRS” and “PRS-CS” demonstrated comparable ability to discriminate between cases and controls, albeit with low accuracy (AAUC [GWAS Hits PRS] = 0.546 [0.490, 0.603], AAUC [PRS-CS] = 0.561 [0.507,0.614]). We also evaluated the performance of a previously published ALS polygenic risk score (PRS-275-non-C9) that excluded the chromosome 9 region and observed limited prediction power (AAUC [PRS-275-non-C9] = 0.539 [0.485, 0.590]) and a relatively weak association with ALS (p-value = 0.10). Since this external PRS didn’t perform better than our two internal PRS and was based on the same ALS GWAS, we did not include this PRS in any additional analyses.

To evaluate if the two risk scores (“GWAS Hits PRS” and “PRS-CS”) capture similar or distinct risk profiles, we combined the two risk scores into the PRS_Combined_, which was evaluated in the same test cohort. As shown in **Table 2**, PRS_Combined_ moderately improved discriminating cases from controls (AAUC [PRS_Combined_]*=* 0.584 [0.525, 0.639]). Additionally, a stronger association of PRS_Combined_ with ALS (P-value = 1.8x10^-5^) was observed than with either individual PRS, suggesting that the joint consideration of oligogenic and polygenic risk profiles can improve ALS prediction models.

**Table 2.** Evaluation of ALS Risk Scores. This table assesses ALS risk scores from genetics (Polygenic Risk Score, PRS), exposures (PXS), and pre-existing conditions (Phenotype Risk Score, PheRS). Scores are evaluated via association (Odds Ratios [OR] with 95% Confidence Intervals [CI] and P-values), accuracy (Brier Score), discrimination (Adjusted Area Under the Curve (AAUC) with 95% CI), and explanatory power (Nagelkerke’s Pseudo-R2). All analyses are adjusted for age, sex, and the first four genetic principal components.

| Predictor |  | Testing Data |  | OR<br>(95% CI) | P value | Brier<br>Score | AAUC<br>(95% CI) | Pseudo<br>-R<sup>2</sup> <sup>a</sup> |
| --- | --- | --- | --- | --- | --- | --- | --- | --- |
|  |  | n<br>Cases | n<br>Controls |  |  |  |  |  |
| Genetics | GWAS Hits<br>PRS<br>(13 variants) | 146 | 1460 | 1.238<br>(1.061, 1.443) | 0.0065 | 0.0822 | 0.533<br>(0.483, 0.582) | 0.0093 |
|  | PRS-CS<br>(1.1M variants) |  |  | 1.210<br>(1.022, 1.433) | 0.027 | 0.0824 | 0.551<br>(0.501, 0.599) | 0.0066 |
|  | GWAS Hits &<br>PRS-CS<br>Combined |  |  | 1.318<br>(1.118, 1.552) | 0.00099 | 0.0820 | 0.568<br>(0.517, 0.617) | 0.014 |
|  | Dou et al. 2023<br>(275 variants) |  |  | 1.150<br>(0.972, 1.360) | 0.10 | 0.0825 | 0.536<br>(0.487, 0.584) | 0.0036 |
| Exposures | PXS | 127 | 1295 | 1.071<br>(0.894, 1.284) | 0.0689 | 0.0813 | 0.518<br>(0.462, 0.571) | 0.0008 |
| Pre-existing<br>conditions /<br>PheRS | >= 0.5 year<br>(PheRS0.5, 8<br>hits) | 110 | 1034 | 1.635<br>(1.416, 1.887) | 2.0E-11 | 0.0810 | 0.572<br>(0.516, 0.628) | 0.081 |
|  | >= 1 year<br>(PheRS1, 5 hits) | 107 | 1012 | 1.433<br>(1.247, 1.647) | 3.9E-07 | 0.0830 | 0.515<br>(0.472, 0.564) | 0.046 |
|  | >= 2 years<br>(PheRS2, 8 hits) | 101 | 979 | 1.091<br>(0.897, 1.328) | 0.38 | 0.0846 | 0.498<br>(0.452, 0.543) | 0.0015 |
|  | >= 3 years<br>(PheRS3, 4 hits) | 96 | 943 | 0.989<br>(0.797, 1.226) | 0.92 | 0.0837 | 0.443<br>(0.416, 0.474) | 2.7E-04 |
|  | >= 4 years<br>(PheRS4, 4 hits) | 92 | 909 | 1.071<br>(0.856, 1.339) | 0.55 | 0.0832 | 0.520<br>(0.483, 0.557) | 8.1E-04 |
|  | >= 5 years<br>(PheRS5, 4 hits) | 86 | 872 | 0.988<br>(0.793, 1.23) | 0.91 | 0.0814 | 0.501<br>(0.462, 0.54) | 1.1E-06 |
| All<br>Categories | Combined PRS | 104 | 994 | 1.303<br>(1.073, 1.582) | 0.0076 | 0.0849 | 0.564<br>(0.506, 0.627) | 0.013 |
|  | PXS |  |  | 1.085<br>(0.889, 1.325) | 0.42 | 0.0855 | 0.522<br>(0.459, 0.579) | 9.4E-04 |
|  | PheRS 1-year<br>threshold |  |  | 1.443<br>(1.255, 1.660) | 2.8E-07 | 0.0821 | 0.518<br>(0.471, 0.565) | 0.049 |
|  | Combined PRS<br>& PheRS1 |  |  | 1.593<br>(1.355, 1.874) | 1.8E-8 | 0.0818 | 0.604<br>(0.547, 0.667) | 0.060 |
|  | All Scores |  |  | 1.437<br>(1.183, 1.747) | 2.7E-04 | 0.0840 | 0.576<br>(0.514, 0.637) | 0.024 |

<sup>a</sup> Nagelkerke [Cragg and Uhler])

### Historical Clinical Data: Phenotype Risk Scores (PheRS)

Next, we explored the influence of pre-existing conditions on ALS risk. We used a PheWAS approach to investigate the enrichment of pre-existing diagnoses in ALS cases versus controls (0.5, 1, 2, 3, 4, or 5 years prior to the onset of ALS, named PheWAS0.5 – PheWAS5, **Figure S3**). In PheWAS0.5, four phecodes, including “Abnormal movement” and “Other acquired deformities of limbs,” reached phenome significance (P-value < 1.4x10^-4^), while only one phecode reached the phenome significance in PheWAS1 (P-value < 1.5x10^-4^). Despite not observing phenome-wide significant associations in the PheWAS2 – PheWAS5, several phenotypes ranked consistently high in almost all the analyses, such as “Pneumonia” and “Other disorders of the circulatory system” (**File S1**). This finding suggests that solely considering phenome-wide significant hits may overlook important factors that might be helpful for prediction purposes.

After identifying these ALS-related phenotype factors, we explored the possibility of predicting ALS by integrating PheWAS results into PheRSs. We expanded our analysis beyond just the phenome-wide significant hits and included the top 50 most significant hits, and performed a forward selection process. The results, shown in **Supplementary Table S1**, indicate that only four to eight phecodes were chosen for each of the six PheWAS. Subsequently, we calculated a PheRS for each PheWAS based on the selected Phecodes and evaluated their predictive performance in the testing cohort (**Table 2)**. Among the six PheRSs evaluated, the PheRS based on symptoms observed 0.5 years before ALS onset exhibited the highest accuracy (AAUC[PheRS0.5] = 0.572 [0.516, 0.628]) while the accuracy of the remaining PheRSs was relatively low. Furthermore, Only PheRS0.5 and PheRS1 showed a strong association with ALS (p-value = 2.0x10^-11^ and 3.9x10^-7^, respectively).

### Health-related Exposure Data: Poly-Exposure Scores (PXS)

We further explored the influence of environmental exposures on ALS development. For this, we relied on a list of 102 potentially relevant exposure variables previously suggested for developing PXS in the UKB study [34]. The evaluation of the PXS in an independent test set of 127 ALS cases and 1,295 controls revealed that it could discriminate cases from controls only with low accuracy (AAUC [PXS] = 0.518 [0.462, 0.571]). Furthermore, the PXS was not significantly associated with ALS (P-value = 0.0689).

### Comparison and Integration of all predictors

In a final step, we integrated PRSs, PheRS, and PXS to improve ALS risk prediction. For this, we opted to include PheRS1 in the combined risk models because it might capture ALS risks earlier even though PheRS0.5 performed better. For completeness, we also included the corresponding results with PheRS0.5 in the supplementary material (**Supplementary Table S4**).

In the first attempt, we constructed a Fully Integrated Risk Score (FIRS) by combining weighted PRSs, PXS, and PheRS1 using the training data. However, the FIRS did not yield satisfactory predictive results on the testing set (AAUC [FIRS] = 0.576 [0.514, 0.637], **Table 2**) and performed worse than the PheRS1 alone in terms of its association with ALS (P-value: 2.7x10^-4^ versus 2.8x10^-7^) and model performance (pseudo-R^2^: 0.024 versus 0.049). Based on the previous results, we hypothesized that this may be due to the limited predictive power of PXS. Consequently, we excluded the PXS and constructed a new composite score only integrating the two PRSs and PheRS1. We called it the Optimal Integrated Risk Score (OIRS) as it exhibited stronger prediction accuracy (AAUC [OIRS] = 0.604 [0.547, 0.667], **Table 2**) than the individual scores and FIRS in the testing set. This improvement is further supported by a stronger association (P-value = 1.8x10^-8^) and the higher pseudo-R^2^ value.

When comparing the distributions of the evaluated scores in ALS cases and controls in the same testing set (n = 104 cases, n = 994 controls, **Figure 3**), we found that OIRS, compared to other scores, exhibited a more pronounced right shift of its distribution in ALS cases versus controls than for any of the individual risk scores or FIRS, which aligned with its superior performance.

**Figure 2.**
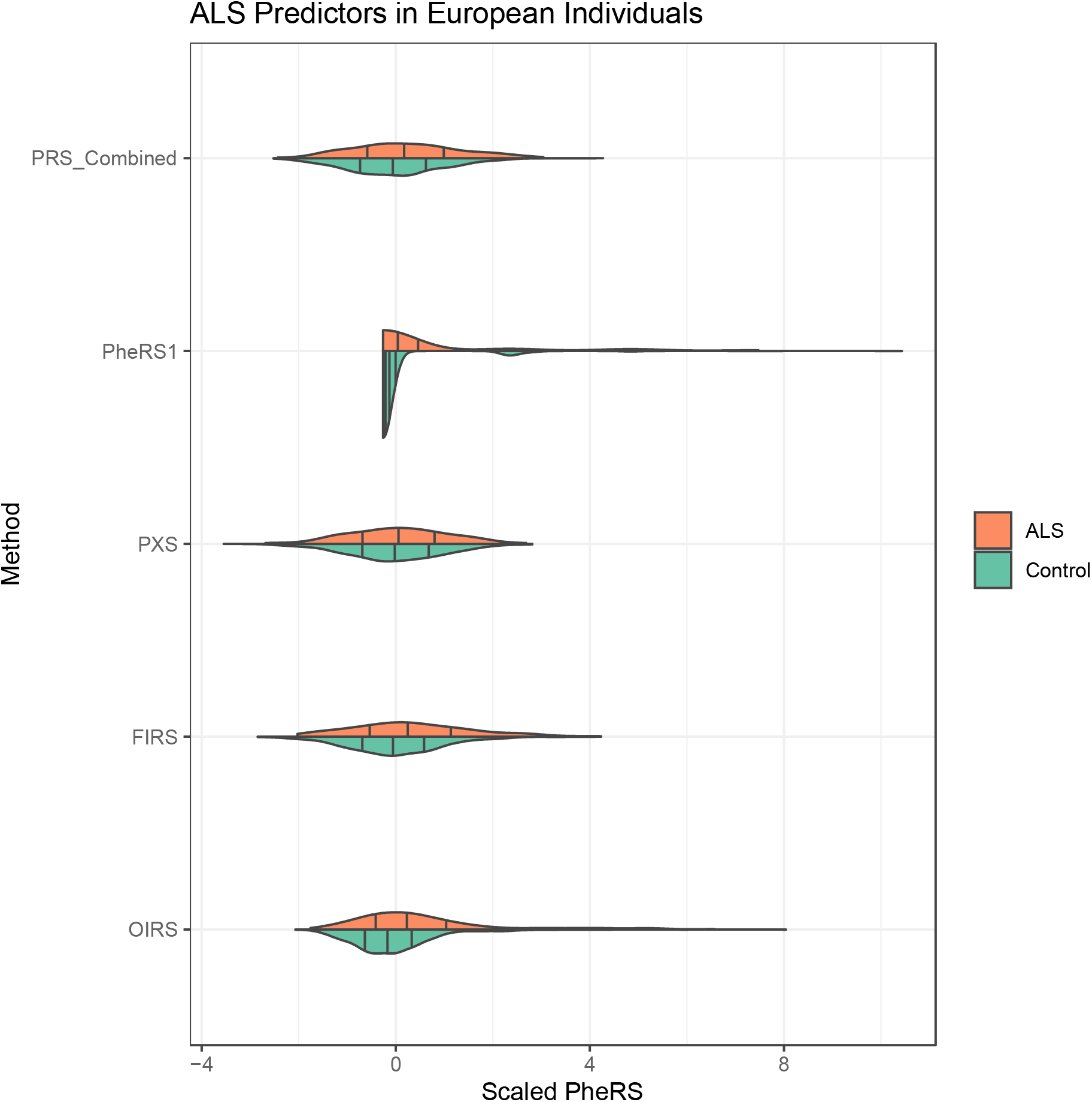
Distribution of ALS Risk Predictors. The violin plots illustrate the distribution of five ALS risk predictors: Combined Polygenic Risk Scores (PRS_Combined_: GWAS Hits -based PRS and PRS-CS based PRS), Poly Exposure Score (PXS), Phenotype Risk Score at 1-year (PheRS1), the Fully Integrated Risk Score (FIRS: both PRS, PXS, and PheRS1), and the Optimal Integrated Risk Score (OIRS: both PRS & PheRS1). Violin plots showcase score distributions for cases (n=104, in orange) and controls (n=994, in green), with black lines indicating the 25th, 50th, and 75th percentile quantiles.

**Figure 3.**
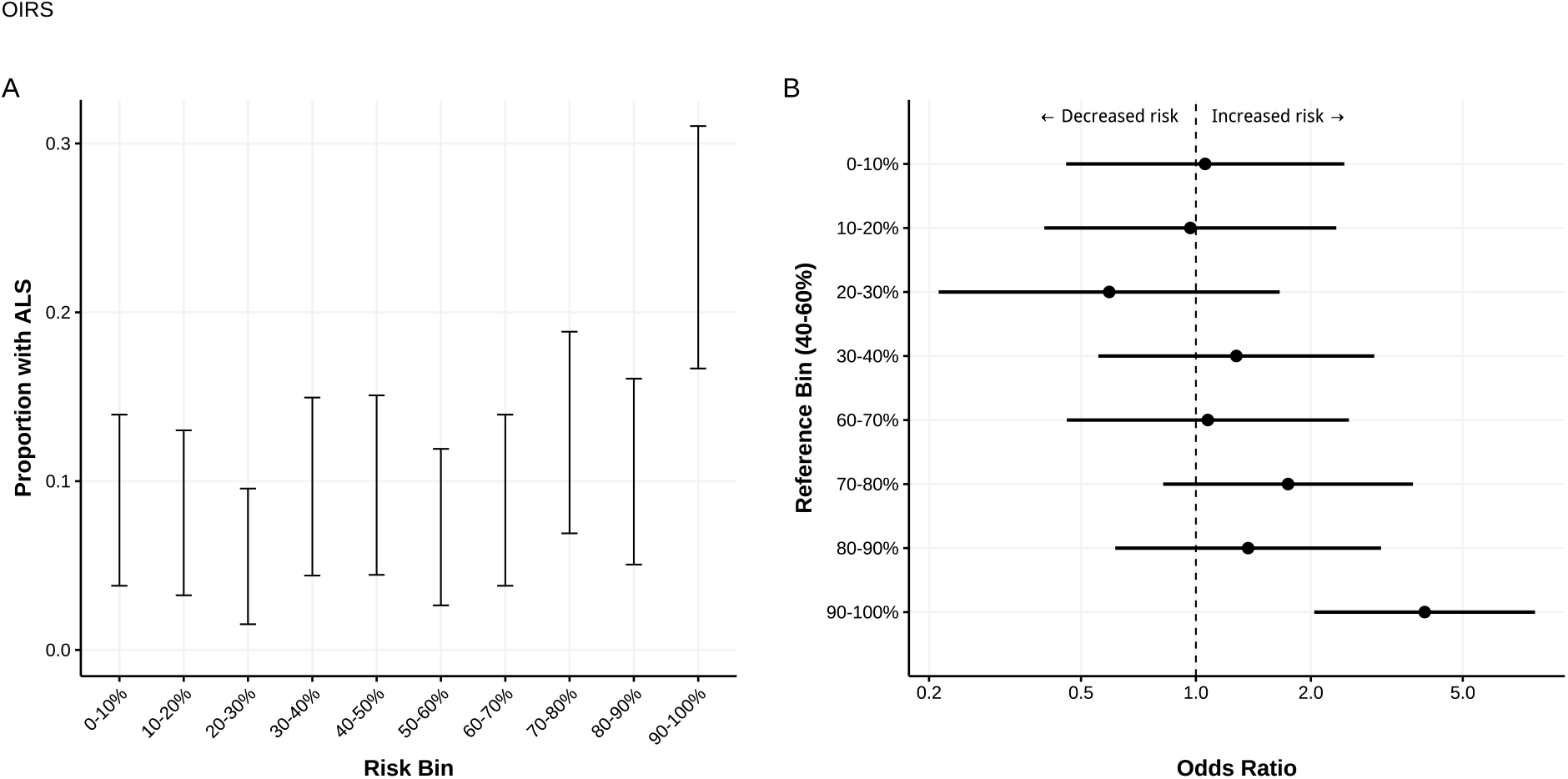
Risk Stratification Based on Optimal Integrated Risk Score (OIRS). This figure shows the risk stratification in the test data (104 cases, 994 controls) using OIRS (integrated from three predictors: GWAS Hits PRS, PRS-CS PRS, and Phenotype Risk Score at 1-year (PheRS1)). **A.** Distribution of ALS cases by OIRS deciles. **B.** ALS Odds Ratio and 95% confidence intervals across aggregate score deciles, with the 40-60% range as the reference group adjusted for sex, PC1 – 4, age, and genotyping array.

Consequently, we adopted OIRS as our final predictor and evaluated its suitability to stratify ALS risk. Although the accuracy of the risk scores was relatively low, indicating the difficulty of predicting ALS at the individual level, we observed that our combined score effectively stratified ALS risk. As shown in **Figure 3A**, OIRS enriched ALS cases in the top 10% risk bin (20.4 % ALS cases) versus the lower nine deciles of the score distribution (3.4 – 12.4%). Specifically, the top 10% of individuals was approximately enriched 4-fold in cases (OR = 3.97 [95% CI: 2.04, 7.73], P-value = 4.9 x 10^-5^) compared to the 40%-60% distribution range (**Figure 3B**, **Supplementary Table S5**), highlighting its potential to identify a subgroup of individuals at substantial risk for ALS.

## Discussion

In this study, we analyzed data from the UK Biobank to develop prediction models for ALS by incorporating various factors from three domains: germline genetics, pre-existing phenotypes, and environmental exposures. We generated PRSs using two different methods, a GWAS hit-based and PRS-CS-based PRS, to capture oligogenic and polygenic risk profiles, respectively. Our PheWAS analyses of various time windows preceding the ALS diagnosis identified several associated pre-existing conditions that we integrated into PheRSs. Additionally, we explored self-reported and environmental exposures, as suggested by He et al. 2021 [34], and generated a PXS. Through systematic assessment of different score combinations, we found the most robust predictor for ALS risk to be the combination of the two PRS and the PheRS1, which aggregated pre-existing diagnoses that occurred at least 1 year before ALS onset. This final prediction model identified a 4-fold increased risk for ALS in the top 10% of the cohort versus the 40%-60% reference, underscoring the potential benefits of integrating a diverse set of associated ALS variables into a comprehensive prediction model.

Building upon this foundation and contextualizing our findings, it is useful to compare them with previous research in the three domains.

*Genetic Risk scores:* We observed significant predictive utility of both our GWAS hits-based PRS and our PRS-CS. Both PRS were based on weights from an independent ALS GWAS. We benchmarked our findings to a previously published ALS PRS, including 275 SNPs (PRS-275-non-C9) [15]. Since this study included the region around C9orf72 to generate the ALS PRSs, but used the same underlying GWAS summary statistics as PRS-275, the poorer performance of the latter may arise because it omitted SNPs from around C9orf72.

*Phenotype risk scores:* The top associations identified in the PheWAS analysis conducted 1 year prior to ALS diagnosis were largely consistent with previous reports. The majority of these associations were musculoskeletal symptoms, such as joint pain [37, 38] and deformities of limbs, and neurological or mental-related symptoms (e.g., abnormal movement [39], neurological disorders [40]). These findings provide evidence of the complex interplay between ALS and circulatory health. While the 1-year window for PheRS may coincide with the average diagnostic delay in ALS, it underscores the potential to forecast the disease a year before the typical diagnosis. Shortening the time to diagnosis, therefore, potentially means patients can get started on therapies sooner when they may be more effective.

*Environmental, diet risk factors / Exposure risk scores*: Furthermore, our study identified several environmental factors associated with the risk of ALS as suggested by the beta coefficients in ridge regression, including the frequency of lamb intake, salt added to food, and stair climbing (**Supplementary Table S6**), consistent with previous works [40-42]. However, the constructed poly environmental score PXS demonstrated weak predictive power. One possible explanation for this low performance is the use of self-reported exposure data, which may introduce inaccuracies. Additionally, the exposures at the time of evaluation may not fully capture the exposures prior to ALS development (On average, the assessment was 5.1 years before ALS.)

Our analysis also demonstrated that the combination of PRSs and PheRS yielded the strongest predictive power for ALS. This finding underscores the importance of considering multiple factors, including genetics and pre-existing phenotypes, in developing accurate ALS prediction models [41]. Further, the ability to predict ALS at the earliest onset of clinical symptoms using a combination of clinical data and genetic data may shorten the time to diagnosis, thereby enabling patients to begin treatment sooner, when they may be more effective.

Our analysis is subject to several limitations that should be acknowledged. Firstly, the lack of a clear definition of ALS in the UK Biobank database led us to use an interchangeable definition for ALS and MND. This broad definition may have included additional, but very rare non-ALS adult MND cases, potentially introducing heterogeneity to our analysis. Secondly, our analysis has limitations regarding overlooking some environmental variables and genetic factors. In our pre-processing methods, we had to exclude 20 environmental variables due to high missing rates and correlations, which might have resulted in omitting potentially influential factors. Additionally, important factors for ALS risk variables, such as persistent organic pollutants [42], were not included in the UK Biobank dataset. Thirdly, our risk scores’ construction relied on regression methods, which may not capture nonlinear combinations of variables influencing ALS prediction[43]. In the future, it may be worthwhile to explore the use of nonlinear methods to improve the accuracy of ALS prediction models [44, 45]. Lastly, our study cohort was derived from the UK Biobank, which primarily represents a European population, and we have limited our analysis to the European ancestry matching the discovery GWAS underlying our PRS [46]. This limits the generalizability of our findings to other populations and reduces our ability to capture the full diversity of ALS risk in the general population.

Our study possesses several notable strengths that contribute to the robustness of our findings. Firstly, we leveraged a large cohort from the UK Biobank, providing a substantial sample size and enabling us to account for potential confounding factors. The comprehensive dataset offered rich genetic, phenotypic, and environmental exposure information, likely enhancing the accuracy of our prediction models by offering a holistic view. Furthermore, we employed various analytical techniques to explore the predictive factors for ALS. By utilizing both univariate regression and ridge regression, we were able to examine the associations between individual variables and ALS risk while effectively managing potential multicollinearity issues. Additionally, we conducted diagnosis-based phenotypic analyses over different time periods, a generalizable approach to explore prediction models that incorporate early symptom onset. Lastly, in our research developed a robust pipeline of methods for disease prediction that incorporates environmental, genetic, and phenotypic factors. This approach is not limited to ALS but has the potential to be applied to other diseases as well.

## Conclusion

There is a critical need to identify risk factors that robustly differentiate ALS cases from healthy controls, facilitating timely ALS risk stratification and early detection. The objective of our study was to identify and investigate pre-disposing factors associated with ALS and to explore them as predictors for ALS. By integrating genetic and phenotypic data, we enhanced our predictive models for ALS.

Looking ahead, there are exciting opportunities to further advance this field. One promising avenue involves incorporating additional, more complex data, encompassing factors like epigenetic factors and biomarkers such as neurofilament light chain and DNA methylation on cytosine bases [47, 48]. This could provide deeper insights into the underlying mechanisms of ALS and enhance the accuracy of predictive models [49]. Additionally, exploring more diverse databases and the potential of nonlinear methods can contribute to a more general and reliable prediction of the disease. The presented framework can also be adapted to explore alternative outcomes, like survival and progression, and by doing so, offer comprehensive insights into the long-term consequences of ALS [50].

## Data Availability

Data cannot be shared publicly due to patient confidentiality. The data underlying the results presented in the study are available from the UK Biobank Platform for researchers who meet the criteria for access to confidential data.

## Web Resources

UK Biobank dataset, https://www.ebi.ac.uk/ega/datasets/EGAD00010001474

PubMed, https://www.ncbi.nlm.nih.gov/pubmed

FRAPOSA, https://github.com/daviddaiweizhang/fraposa

R package “Rprs”, https://github.com/statgen/Rprs

Weights for constructed PXS and PRS, https://csg.sph.umich.edu/larsf/SuppData/ALS_UKB_2023/

Definitions of Motor Neurone Disease and the Major Diagnostic Subtypes, https://biobank.ndph.ox.ac.uk/showcase/showcase/docs/alg_outcome_mnd.pdf

## Author Contributions

W.J.: writing—original draft, writing—review and editing, visualization. A.J.A.: writing—review and editing. J.B.: writing—review and editing. K.M.B.: writing—review and editing. S.A.G.: writing—review and editing. E.L.F.: writing—review and editing. B.M.: conceptualization, methodology, writing—original draft, writing—review and editing, supervision, funding acquisition. L.G.F.: conceptualization, methodology, formal analysis, investigation, data curation, writing—original draft, writing—review and editing, visualization. All authors have read and agreed to the published version of the manuscript.

## Acknowledgment

This research has been conducted using the UK Biobank Resource under application number 24460. KMB and SAG received funding through the ALS Association (20-IIA-532). SAG and ELF received funding through the CDC (R01TS000344) and NIH (R01NS127188, R01ES030049). SAG received funding through the NIH (K23ES027221).

