## Supplement Tables and Figures for "Improving prediction models of amyotrophic lateral sclerosis (ALS) using polygenic, pre-existing conditions, and survey-based risk scores in the UK Biobank"

### Supplementary Tables

**Table S1** Weights in Firth-corrected logistic regression after forward selection using Phenome with different time thresholds. Not all 50 Phecodes were presented in the table. "Phecode" column contains the numerical value of the Phecode, while the "Phecode Description" column provides a description of the corresponding Phecode. The "Category" column indicates the diagnosis category to which the Phecode belongs. The "Year before ALS onset" column specifies the time threshold of the Phenome used for forward selection. The numeric values in the table represent the beta coefficient of each Phecode in the regression (also serve as the weight for calculating Phenotype Risk Scores [PheRS]). Phecodes that were dropped during the forward selection process are denoted by "..." in the table.

| **Phecode** | **Phecode Description** | **Category** | **Beta Coefficient / PheRS Weight** | | | | | |
| --- | --- | --- | --- | --- | --- | --- | --- | --- |
|  |  |  | **Year before ALS onset** | | | | | |
|  |  |  | **0.5** | **1** | **2** | **3** | **4** | **5** |
| 276.1 | Electrolyte Imbalance | Endocrine/Metabolic | ... | ... | 1.699 | ... | ... | ... |
| 292.1 | Aphasia/Speech Disturbance | Mental Disorders | 1.95 | ... | ... | ... | ... | ... |
| 350 | Abnormal Movement | Neurological | 2.339 | 2.675 | ... | ... | ... | ... |
| 361 | Retinal Detachments and Defects | Sense Organs | ... | ... | ... | ... | 1.461 | ... |
| 433.2 | Occlusion of Cerebral Arteries | Circulatory System | 1.378 | ... | ... | 1.519 | ... | ... |
| 443 | Peripheral Vascular Disease | Circulatory System | ... | ... | ... | 1.378 | 1.472 | ... |
| 459 | Other Disorders of Circulatory System | Circulatory System | ... | ... | ... | ... | ... | 1.184 |
| 480 | Pneumonia | Respiratory | 1.11 | 1.388 | 1.156 | ... | ... | ... |
| 530.14 | Reflux Esophagitis | Digestive | ... | ... | 1.085 | ... | ... | ... |
| 535 | Gastritis and Duodenitis | Digestive | ... | ... | -1.383 | ... | ... | ... |
| 537 | Other Disorders of Stomach and Duodenum | Digestive | ... | ... | 1.443 | 1.559 | 1.341 | ... |
| 550.4 | Umbilical Hernia | Digestive | 1.403 | ... | ... | ... | ... | 1.464 |
| 569 | Other Disorders of Intestine | Digestive | ... | ... | ... | ... | -2.111 | -2.024 |
| 720 | Spinal Stenosis | Musculoskeletal | ... | ... | 1.419 | ... | ... | ... |
| 736 | Other Acquired Deformities of Limbs | Musculoskeletal | 1.878 | 1.602 | 1.595 | 1.66 | ... | ... |
| 742.9 | Other Derangement of Joint | Musculoskeletal | 1.541 | 1.424 | 1.293 | ... | ... | 1.4 |
| 781 | Symptoms Involving Nervous and Musculoskeletal Systems | Symptoms | 1.418 | 1.367 | ... | ... | ... | ... |

**Table S2** Statistics of exposure missingness when creating the Poly Exposure Score in overall (or ALS cases, controls) cohort. The “Variable” column lists the exposure variables included in the analysis. The "Missingness" column represents the number of missing entries and the percentage of missingness in the overall cohort (or ALS cases, controls). The odds ratio and p-value represent Fisher’s Exact test results when comparing the missingness between cases and controls. Exposures were excluded from the analysis if the overall missingness exceeded 20% (highlighted in red).

| **Variable** | **Category** | **Missing Entries** | | | | | | **Missing in ALS vs. Matched Controls (Fisher’s Exact Test)** | |
| --- | --- | --- | --- | --- | --- | --- | --- | --- | --- |
|  |  | **Overall n = 3212** | | **ALS n = 292** | | **Matched Controls n = 2920** | |  |  |
|  |  | **n Miss.** | **%** | **n Miss.** | **%** | **n Miss.** | **%** | **Odds Ratio** | **P** |
| Alcohol intake frequency. | Alcohol | 4 | 0.12 | 1 | 0.34 | 3 | 0.1 | 3.34 (0.0634, 41.7) | 0.317 |
| Alcohol intake versus 10 years previously | Alcohol | 238 | 7.41 | 26 | 8.9 | 212 | 7.26 | 1.25 (0.782, 1.92) | 0.293 |
| Alcohol drinker status | Alcohol | 5 | 0.16 | 1 | 0.34 | 4 | 0.14 | 2.5 (0.0507, 25.4) | 0.379 |
| Townsend deprivation index at recruitment | Baseline | 9 | 0.28 | 2 | 0.68 | 7 | 0.24 | 2.87 (0.289, 15.2) | 0.194 |
| Cooked vegetable intake | Diet | 37 | 1.15 | 6 | 2.05 | 31 | 1.06 | 1.95 (0.661, 4.81) | 0.143 |
| Salad / raw vegetable intake | Diet | 43 | 1.34 | 6 | 2.05 | 37 | 1.27 | 1.63 (0.559, 3.95) | 0.278 |
| Fresh fruit intake | Diet | 13 | 0.4 | 1 | 0.34 | 12 | 0.41 | 0.833 (0.0194, 5.67) | 1 |
| Dried fruit intake | Diet | 38 | 1.18 | 3 | 1.03 | 35 | 1.2 | 0.856 (0.167, 2.74) | 1 |
| Oily fish intake | Diet | 19 | 0.59 | 1 | 0.34 | 18 | 0.62 | 0.554 (0.0133, 3.53) | 1 |
| Non-oily fish intake | Diet | 9 | 0.28 | 1 | 0.34 | 8 | 0.27 | 1.25 (0.0281, 9.38) | 0.576 |
| Processed meat intake | Diet | 7 | 0.22 | 1 | 0.34 | 6 | 0.21 | 1.67 (0.0362, 13.8) | 0.487 |
| Poultry intake | Diet | 10 | 0.31 | 2 | 0.68 | 8 | 0.27 | 2.51 (0.258, 12.7) | 0.229 |
| Beef intake | Diet | 9 | 0.28 | 1 | 0.34 | 8 | 0.27 | 1.25 (0.0281, 9.38) | 0.576 |
| Lamb/mutton intake | Diet | 19 | 0.59 | 2 | 0.68 | 17 | 0.58 | 1.18 (0.131, 5) | 0.689 |
| Pork intake | Diet | 17 | 0.53 | 3 | 1.03 | 14 | 0.48 | 2.15 (0.395, 7.78) | 0.196 |
| Cheese intake | Diet | 91 | 2.83 | 12 | 4.11 | 79 | 2.71 | 1.54 (0.755, 2.89) | 0.191 |
| Milk type used | Diet | 0 | 0 | 0 | 0 | 0 | 0 | n/a | 1 |
| Spread type | Diet | 3 | 0.09 | 1 | 0.34 | 2 | 0.07 | 5.01 (0.0847, 96.6) | 0.249 |
| Bread intake | Diet | 38 | 1.18 | 8 | 2.74 | 30 | 1.03 | 2.71 (1.06, 6.13) | 0.0185 |
| Bread type | Diet | 93 | 2.9 | 8 | 2.74 | 85 | 2.91 | 0.94 (0.389, 1.96) | 1 |
| Cereal intake | Diet | 5 | 0.16 | 1 | 0.34 | 4 | 0.14 | 2.5 (0.0507, 25.4) | 0.379 |
| Salt added to food | Diet | 1 | 0.03 | 0 | 0 | 1 | 0.03 | 0 (0, 388) | 1 |
| Tea intake | Diet | 8 | 0.25 | 1 | 0.34 | 7 | 0.24 | 1.43 (0.0316, 11.2) | 0.534 |
| Coffee intake | Diet | 7 | 0.22 | 0 | 0 | 7 | 0.24 | 0 (0, 6.96) | 1 |
| Hot drink temperature | Diet | 17 | 0.53 | 1 | 0.34 | 16 | 0.55 | 0.624 (0.0148, 4.04) | 1 |
| Water intake | Diet | 29 | 0.9 | 3 | 1.03 | 26 | 0.89 | 1.16 (0.222, 3.81) | 0.743 |
| Major dietary changes in the last 5 years | Diet | 3 | 0.09 | 0 | 0 | 3 | 0.1 | 0 (0, 24.2) | 1 |
| Variation in diet | Diet | 8 | 0.25 | 1 | 0.34 | 7 | 0.24 | 1.43 (0.0316, 11.2) | 0.534 |
| Never eat eggs, dairy, wheat, sugar | Diet | 5 | 0.16 | 1 | 0.34 | 4 | 0.14 | 2.5 (0.0507, 25.4) | 0.379 |
| Beef intake (2) | Meat/fish yesterday | 2777 | 86.46 | 262 | 89.73 | 2515 | 86.13 |  |  |
| Pork intake (2) | Meat/fish yesterday | 2998 | 93.34 | 277 | 94.86 | 2721 | 93.18 | 1.41 (0.946, 2.16) | 0.0888 |
| Poultry intake (2) | Meat/fish yesterday | 2710 | 84.37 | 257 | 88.01 | 2453 | 84.01 | 1.35 (0.785, 2.49) | 0.325 |
| Oily fish intake (2) | Meat/fish yesterday | 2919 | 90.88 | 268 | 91.78 | 2651 | 90.79 | 1.4 (0.964, 2.08) | 0.0759 |
| Birth weight | Early life factors | 1564 | 48.69 | 145 | 49.66 | 1419 | 48.6 | 1.04 (0.814, 1.34) | 0.759 |
| Country of birth (UK/elsewhere) | Early life factors | 1 | 0.03 | 0 | 0 | 1 | 0.03 | 0 (0, 388) | 1 |
| Breastfed as a baby | Early life factors | 896 | 27.9 | 88 | 30.14 | 808 | 27.67 | 1.13 (0.857, 1.48) | 0.374 |
| Comparative body size at age 10 | Early life factors | 58 | 1.81 | 6 | 2.05 | 52 | 1.78 | 1.16 (0.403, 2.73) | 0.647 |
| Comparative height size at age 10 | Early life factors | 51 | 1.59 | 5 | 1.71 | 46 | 1.58 | 1.09 (0.335, 2.76) | 0.805 |
| Handedness (chirality/laterality) | Early life factors | 2 | 0.06 | 0 | 0 | 2 | 0.07 | 0 (0, 53.3) | 1 |
| Adopted as a child | Early life factors | 4 | 0.12 | 3 | 1.03 | 1 | 0.03 | 30.2 (2.42, 1570) | 0.00278 |
| Part of a multiple birth | Early life factors | 50 | 1.56 | 4 | 1.37 | 46 | 1.58 | 0.868 (0.225, 2.4) | 1 |
| Maternal smoking around birth | Early life factors | 505 | 15.72 | 45 | 15.41 | 460 | 15.75 | 0.974 (0.682, 1.37) | 0.933 |
| Qualifications | Education | 31 | 0.97 | 4 | 1.37 | 27 | 0.92 | 1.49 (0.376, 4.31) | 0.522 |
| Current employment status | Employment | 27 | 0.84 | 2 | 0.68 | 25 | 0.86 | 0.799 (0.0912, 3.23) | 1 |
| Type of accommodation lived in | Household | 5 | 0.16 | 1 | 0.34 | 4 | 0.14 | 2.5 (0.0507, 25.4) | 0.379 |
| Own or rent accommodation lived in | Household | 55 | 1.71 | 9 | 3.08 | 46 | 1.58 | 1.99 (0.846, 4.16) | 0.0906 |
| Length of time at current address | Household | 6 | 0.19 | 0 | 0 | 6 | 0.21 | 0 (0, 8.52) | 1 |
| Number in household | Household | 30 | 0.93 | 7 | 2.4 | 23 | 0.79 | 3.09 (1.11, 7.52) | 0.0156 |
| Number of vehicles in household | Household | 32 | 1 | 6 | 2.05 | 26 | 0.89 | 2.33 (0.779, 5.86) | 0.0646 |
| Average total household income before tax | Household | 491 | 15.29 | 53 | 18.15 | 438 | 15 | 1.26 (0.899, 1.73) | 0.172 |
| Gas or solid-fuel cooking/heating | Household | 3 | 0.09 | 2 | 0.68 | 1 | 0.03 | 20.1 (1.04, 1180) | 0.0232 |
| Number of days/week walked 10+ minutes | Physical activity | 52 | 1.62 | 7 | 2.4 | 45 | 1.54 | 1.57 (0.591, 3.55) | 0.324 |
| Duration of walks | Physical activity | 422 | 13.14 | 45 | 15.41 | 377 | 12.91 | 1.23 (0.858, 1.73) | 0.237 |
| Number of days/week of moderate physical activity 10+ minutes | Physical activity | 170 | 5.29 | 20 | 6.85 | 150 | 5.14 | 1.36 (0.793, 2.22) | 0.217 |
| Number of days/week of vigorous physical activity 10+ minutes | Physical activity | 168 | 5.23 | 18 | 6.16 | 150 | 5.14 | 1.21 (0.689, 2.02) | 0.411 |
| Usual walking pace | Physical activity | 20 | 0.62 | 5 | 1.71 | 15 | 0.51 | 3.37 (0.952, 9.85) | 0.0297 |
| Frequency of stair climbing in last 4 weeks | Physical activity | 29 | 0.9 | 4 | 1.37 | 25 | 0.86 | 1.61 (0.404, 4.7) | 0.33 |
| Time spent watching television (TV) | Physical activity | 27 | 0.84 | 5 | 1.71 | 22 | 0.75 | 2.29 (0.674, 6.27) | 0.0922 |
| Time spent using computer | Physical activity | 24 | 0.75 | 0 | 0 | 24 | 0.82 | 0 (0, 1.66) | 0.161 |
| Time spent driving | Physical activity | 50 | 1.56 | 8 | 2.74 | 42 | 1.44 | 1.93 (0.775, 4.21) | 0.129 |
| Drive faster than motorway speed limit | Physical activity | 88 | 2.74 | 14 | 4.79 | 74 | 2.53 | 1.94 (0.996, 3.51) | 0.0358 |
| Types of transport used (excluding work) | Physical activity | 14 | 0.44 | 2 | 0.68 | 12 | 0.41 | 1.67 (0.181, 7.56) | 0.368 |
| Types of physical activity in last 4 weeks | Physical activity | 13 | 0.4 | 2 | 0.68 | 11 | 0.38 | 1.82 (0.195, 8.41) | 0.334 |
| Home area population density - urban or rural | Reception | 33 | 1.03 | 5 | 1.71 | 28 | 0.96 | 1.8 (0.538, 4.77) | 0.218 |
| Nitrogen dioxide air pollution; 2010 | Residential noise pollution | 42 | 1.31 | 3 | 1.03 | 39 | 1.34 | 0.767 (0.151, 2.44) | 1 |
| Nitrogen oxides air pollution; 2010 | Residential noise pollution | 42 | 1.31 | 3 | 1.03 | 39 | 1.34 | 0.767 (0.151, 2.44) | 1 |
| Particulate matter air pollution (pm10); 2010 | Residential noise pollution | 281 | 8.75 | 24 | 8.22 | 257 | 8.8 | 0.928 (0.573, 1.44) | 0.828 |
| Particulate matter air pollution (pm2.5); 2010 | Residential noise pollution | 281 | 8.75 | 24 | 8.22 | 257 | 8.8 | 0.928 (0.573, 1.44) | 0.828 |
| Particulate matter air pollution (pm2.5) absorbance; 2010 | Residential noise pollution | 281 | 8.75 | 24 | 8.22 | 257 | 8.8 | 0.928 (0.573, 1.44) | 0.828 |
| Particulate matter air pollution 2.5-10um; 2010 | Residential noise pollution | 281 | 8.75 | 24 | 8.22 | 257 | 8.8 | 0.928 (0.573, 1.44) | 0.828 |
| Traffic intensity on the nearest road | Residential noise pollution | 42 | 1.31 | 3 | 1.03 | 39 | 1.34 | 0.767 (0.151, 2.44) | 1 |
| Inverse distance to the nearest road | Residential noise pollution | 42 | 1.31 | 3 | 1.03 | 39 | 1.34 | 0.767 (0.151, 2.44) | 1 |
| Traffic intensity on the nearest major road | Residential noise pollution | 42 | 1.31 | 3 | 1.03 | 39 | 1.34 | 0.767 (0.151, 2.44) | 1 |
| Inverse distance to the nearest major road | Residential noise pollution | 42 | 1.31 | 3 | 1.03 | 39 | 1.34 | 0.767 (0.151, 2.44) | 1 |
| Total traffic load on major roads | Residential noise pollution | 42 | 1.31 | 3 | 1.03 | 39 | 1.34 | 0.767 (0.151, 2.44) | 1 |
| Close to major road | Residential noise pollution | 42 | 1.31 | 3 | 1.03 | 39 | 1.34 | 0.767 (0.151, 2.44) | 1 |
| Sum of road length of major roads within 100m | Residential noise pollution | 42 | 1.31 | 3 | 1.03 | 39 | 1.34 | 0.767 (0.151, 2.44) | 1 |
| Nitrogen dioxide air pollution; 2005 | Residential noise pollution | 42 | 1.31 | 3 | 1.03 | 39 | 1.34 | 0.767 (0.151, 2.44) | 1 |
| Nitrogen dioxide air pollution; 2006 | Residential noise pollution | 42 | 1.31 | 3 | 1.03 | 39 | 1.34 | 0.767 (0.151, 2.44) | 1 |
| Nitrogen dioxide air pollution; 2007 | Residential noise pollution | 42 | 1.31 | 3 | 1.03 | 39 | 1.34 | 0.767 (0.151, 2.44) | 1 |
| Particulate matter air pollution (pm10); 2007 | Residential noise pollution | 49 | 1.53 | 3 | 1.03 | 46 | 1.58 | 0.649 (0.128, 2.04) | 0.62 |
| Average daytime sound level of noise pollution | Residential noise pollution | 42 | 1.31 | 3 | 1.03 | 39 | 1.34 | 0.767 (0.151, 2.44) | 1 |
| Average evening sound level of noise pollution | Residential noise pollution | 42 | 1.31 | 3 | 1.03 | 39 | 1.34 | 0.767 (0.151, 2.44) | 1 |
| Average night-time sound level of noise pollution | Residential noise pollution | 42 | 1.31 | 3 | 1.03 | 39 | 1.34 | 0.767 (0.151, 2.44) | 1 |
| Average 16-hour sound level of noise pollution | Residential noise pollution | 42 | 1.31 | 3 | 1.03 | 39 | 1.34 | 0.767 (0.151, 2.44) | 1 |
| Average 24-hour sound level of noise pollution | Residential noise pollution | 42 | 1.31 | 3 | 1.03 | 39 | 1.34 | 0.767 (0.151, 2.44) | 1 |
| Age first had sexual intercourse | Sexual factors | 379 | 11.8 | 32 | 10.96 | 347 | 11.88 | 0.913 (0.601, 1.35) | 0.704 |
| Sleep duration | Sleep | 17 | 0.53 | 2 | 0.68 | 15 | 0.51 | 1.34 (0.148, 5.79) | 0.663 |
| Getting up in morning | Sleep | 3 | 0.09 | 3 | 1.03 | 0 | 0 | Inf (4.15, Inf) | 0.000744 |
| Morning/evening person (chronotype) | Sleep | 352 | 10.96 | 35 | 11.99 | 317 | 10.86 | 1.12 (0.748, 1.63) | 0.555 |
| Nap during day | Sleep | 1 | 0.03 | 0 | 0 | 1 | 0.03 | 0 (0, 388) | 1 |
| Sleeplessness / insomnia | Sleep | 3 | 0.09 | 0 | 0 | 3 | 0.1 | 0 (0, 24.2) | 1 |
| Snoring | Sleep | 219 | 6.82 | 25 | 8.56 | 194 | 6.64 | 1.32 (0.815, 2.05) | 0.223 |
| Daytime dozing / sleeping (narcolepsy) | Sleep | 9 | 0.28 | 1 | 0.34 | 8 | 0.27 | 1.25 (0.0281, 9.38) | 0.576 |
| Current tobacco smoking | Smoking | 3 | 0.09 | 0 | 0 | 3 | 0.1 | 0 (0, 24.2) | 1 |
| Past tobacco smoking | Smoking | 245 | 7.63 | 26 | 8.9 | 219 | 7.5 | 1.21 (0.755, 1.86) | 0.417 |
| Smoking/smokers in household | Smoking | 234 | 7.29 | 24 | 8.22 | 210 | 7.19 | 1.16 (0.711, 1.81) | 0.481 |
| Exposure to tobacco smoke at home | Smoking | 295 | 9.18 | 29 | 9.93 | 266 | 9.11 | 1.1 (0.708, 1.66) | 0.67 |
| Exposure to tobacco smoke outside home | Smoking | 485 | 15.1 | 55 | 18.84 | 430 | 14.73 | 1.34 (0.966, 1.84) | 0.0714 |
| Smoking status | Smoking | 12 | 0.37 | 2 | 0.68 | 10 | 0.34 | 2.01 (0.213, 9.48) | 0.299 |
| Ever smoked | Smoking | 13 | 0.4 | 2 | 0.68 | 11 | 0.38 | 1.82 (0.195, 8.41) | 0.334 |

**Table S3** Variable statistics for VIF (variance inflation factor) filtering among 50 imputations. Variables with more than 25 high VIFs among 50 imputations were excluded from the analysis.

| **Variable** | **n Observations with VIF > 5** |
| --- | --- |
| Alcohol Intake Frequency | 50 |
| Average 24-Hour Sound Level of Noise Pollution | 50 |
| Average Daytime Sound Level of Noise Pollution | 50 |
| Average Evening Sound Level of Noise Pollution | 50 |
| Current Employment Status | 50 |
| Nitrogen Dioxide Air Pollution 2005 | 50 |
| Nitrogen Dioxide Air Pollution 2007 | 50 |
| Nitrogen Dioxide Air Pollution 2010 | 50 |
| Nitrogen Oxides Air Pollution 2010 | 50 |
| Particulate Matter Air Pollution 2.5-10µm 2010 | 50 |
| Past Tobacco Smoking | 50 |
| Sum of Road Length of Major Roads Within 100m | 50 |
| Average 16-Hour Sound Level of Noise Pollution | 36 |
| Average Nighttime Sound Level of Noise Pollution | 14 |
| Nitrogen Dioxide Air Pollution 2006 | 4 |
| Average Total Household Income Before Tax | 0 |
| Frequency of Stair Climbing in Last 4 Weeks | 0 |
| Inverse Distance to the Nearest Major Road | 0 |
| Milk Type Used | 0 |
| Number of Vehicles in Household | 0 |
| Own or Rent Accommodation Lived In | 0 |
| Particulate Matter Air Pollution PM10 2010 | 0 |
| Qualifications | 0 |
| Total Traffic Load on Major Roads | 0 |
| Types of Physical Activity in Last 4 Weeks | 0 |

**Table S4** Evaluation of ALS Risk Scores. This table assesses the ALS risk scores from genetics (Polygenic Risk Score, PRS), exposures (PXS), and pre-existing conditions (Phenotype Risk Score, PheRS constructed based on diagnosis that were half a year prior to the onset of ALS) on the same cohort.

Scores are evaluated via association (Odds Ratios [OR] with 95% Confidence Intervals [CI] and P-values), accuracy (Brier Score), discrimination (Adjusted Area Under the Curve (AAUC) with 95% CI), and explanatory power (Nagelkerke’s Pseudo-R2). All analyses are adjusted for age, sex, and the first four genetic principal components.

| **Predictor** | | **Testing Data** | | **OR (95% CI)^a^** | **P value** | **Brier Score** | **AAUC (95% CI)** | **Pseudo-R2 ^a^** |
| --- | --- | --- | --- | --- | --- | --- | --- | --- |
|  |  | **n Cases** | **n Controls** |  |  |  |  |  |
| All Categories | PRS_Combined_ | 107 | 1016 | 1.303 (1.076, 1.578) | 0.0066 | 0.0853 | 0.562 (0.500, 0.621) | 0.013 |
|  | PXS |  |  | 1.081 (0.887, 1.319) | 0.44 | 0.0859 | 0.520 (0.462, 0.58) | 9.0E-04 |
|  | PheRS 0.5-year threshold |  |  | 1.654 (1.431, 1.913) | 1.0E-11 | 0.0800 | 0.578 (0.524, 0.633) | 0.086 |
|  | PRS_Combined_ & PheRS0.5 |  |  | 1.807 (1.535, 2.127) | 1.1E-12 | 0.0797 | 0.637 (0.577, 0.695) | 0.094 |
|  | PRS_Combined_, PXS and PheRS0.5) |  |  | 1.652 (1.367, 1.995) | 2.0E-7 | 0.0829 | 0.613 (0.554, 0.674) | 0.048 |

^a^ Nagelkerke [Cragg and Uhler])

**Table S5. Risk Stratification Using the Optimal Integrated Risk Score (OIRS).** The figure presents the Odds Ratio for ALS and its associated 95% confidence intervals (CI) across OIRS deciles. The 40-60% range serves as the reference group. Results are adjusted for sex, the first four principal components (PC1-4), age, and genotyping array. Association p-values are also displayed.

| **Risk Bin** | **ALS Odds Ratio**  **(95% CI)** | **P-value** |
| --- | --- | --- |
| 0-10% | 1.057 (0.457, 2.447) | 0.90 |
| 10-20% | 0.967 (0.401, 2.331 | 0.94 |
| 20-30% | 0.593 (0.212, 1.657) | 0.32 |
| 30-40% | 1.276 (0.555, 2.932) | 0.57 |
| 60-70% | 1.074 (0.459, 2.516) | 0.87 |
| 70-80% | 1.743 (0.821, 3.701) | 0.15 |
| 80-90% | 1.370 (0.615, 3.055) | 0.44 |
| 90-100% | 3.973 (2.042, 7.73) | 4.90 x 10^-5^ |

### Supplementary Figures

**Figure S1** Sample Selection and Filtering Flowchart. The sequential selection process for ALS cases (orange) and controls (light green) is shown. Grey boxes indicate exclusion steps due to specific criteria. The flow begins with data sources, moves through ALS case identification, genotype data cleaning, and culminates in the control matching procedure with training testing data split.

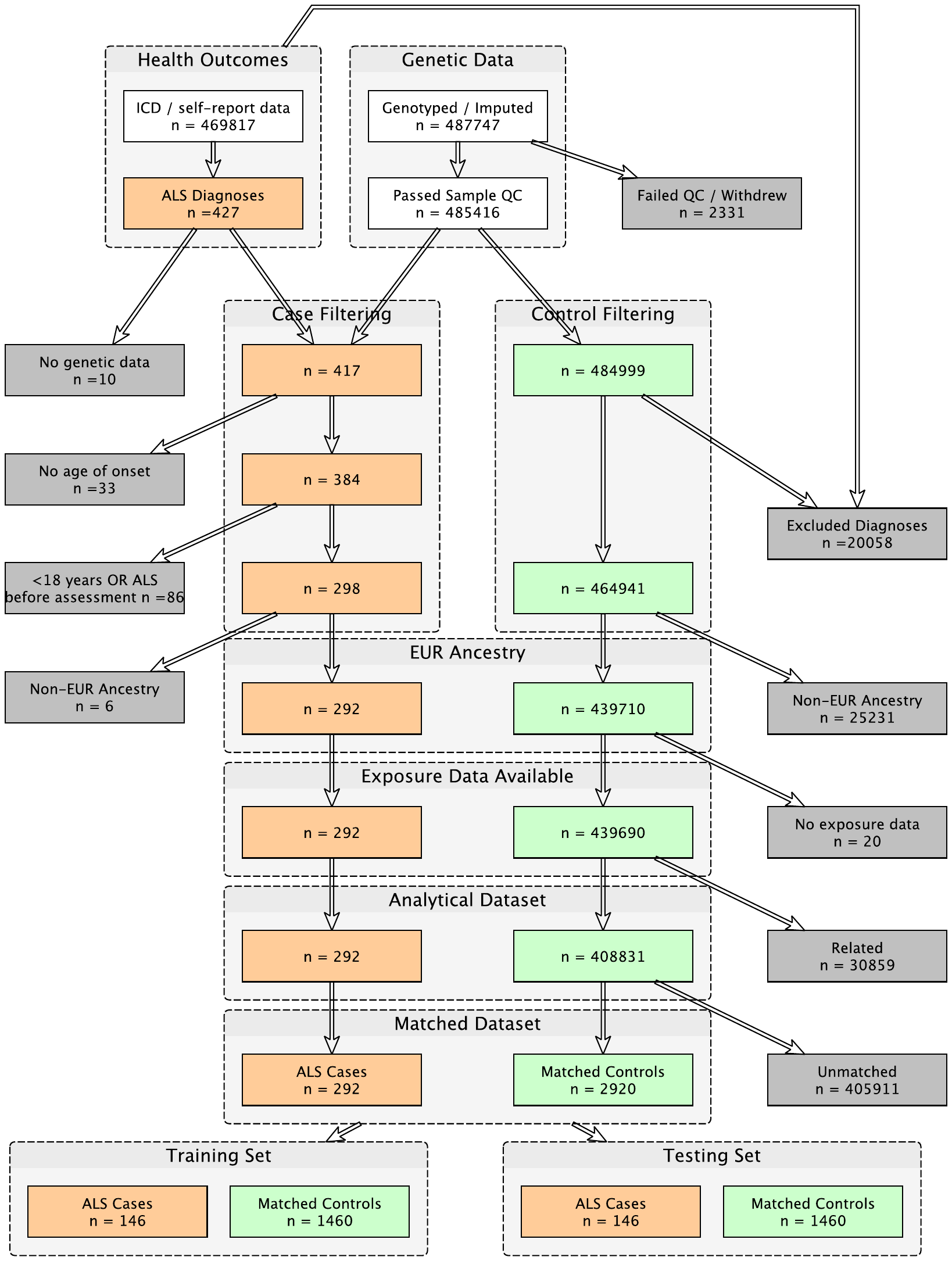

**Figure S2** Age distribution at first motor neuron disease (ALS) diagnosis among 392 ALS cases, stratified by sex. Female and male are labeled as red and blue.

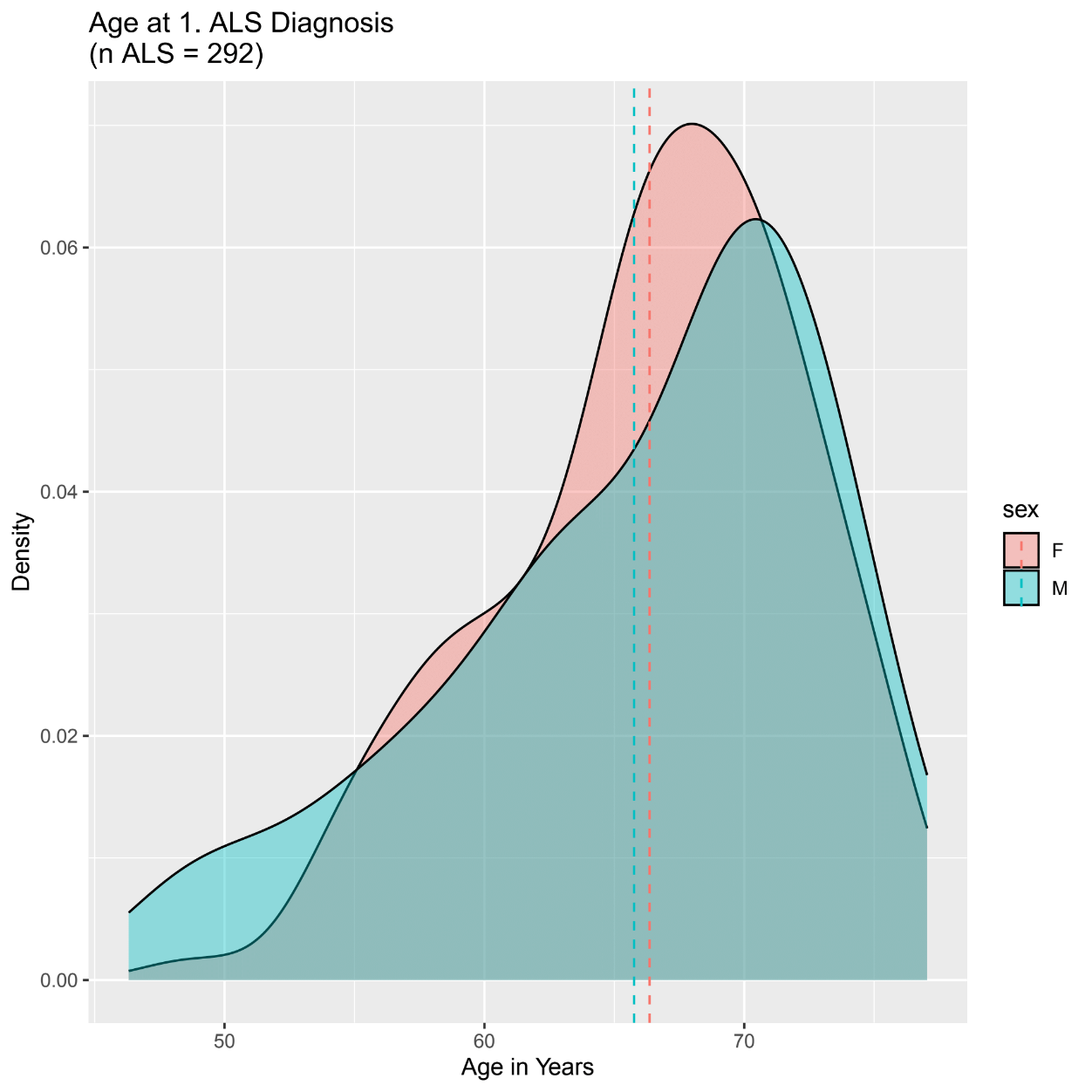

**Figure S3** PheWAS on Pre-ALS Symptoms. This figure presents a PheWAS on symptoms appearing at least A: 0.5 year; B: 1 years; C: 2 years; D: 3 years; E: 4 years; F: 5 years before ALS onset. The number of ALS cases, controls, and phecodes were labeled on top of the figures. The analysis was adjusted for age, sex, and four principal components. Phenome-wide (red) and nominal (orange) significance are indicated as horizontal dotted lines. The top ten associated traits are labeled, while directional triangles show positive (upwards) or negative (downwards) association with ALS outcome. he horizontal dotted lines mean

**A**

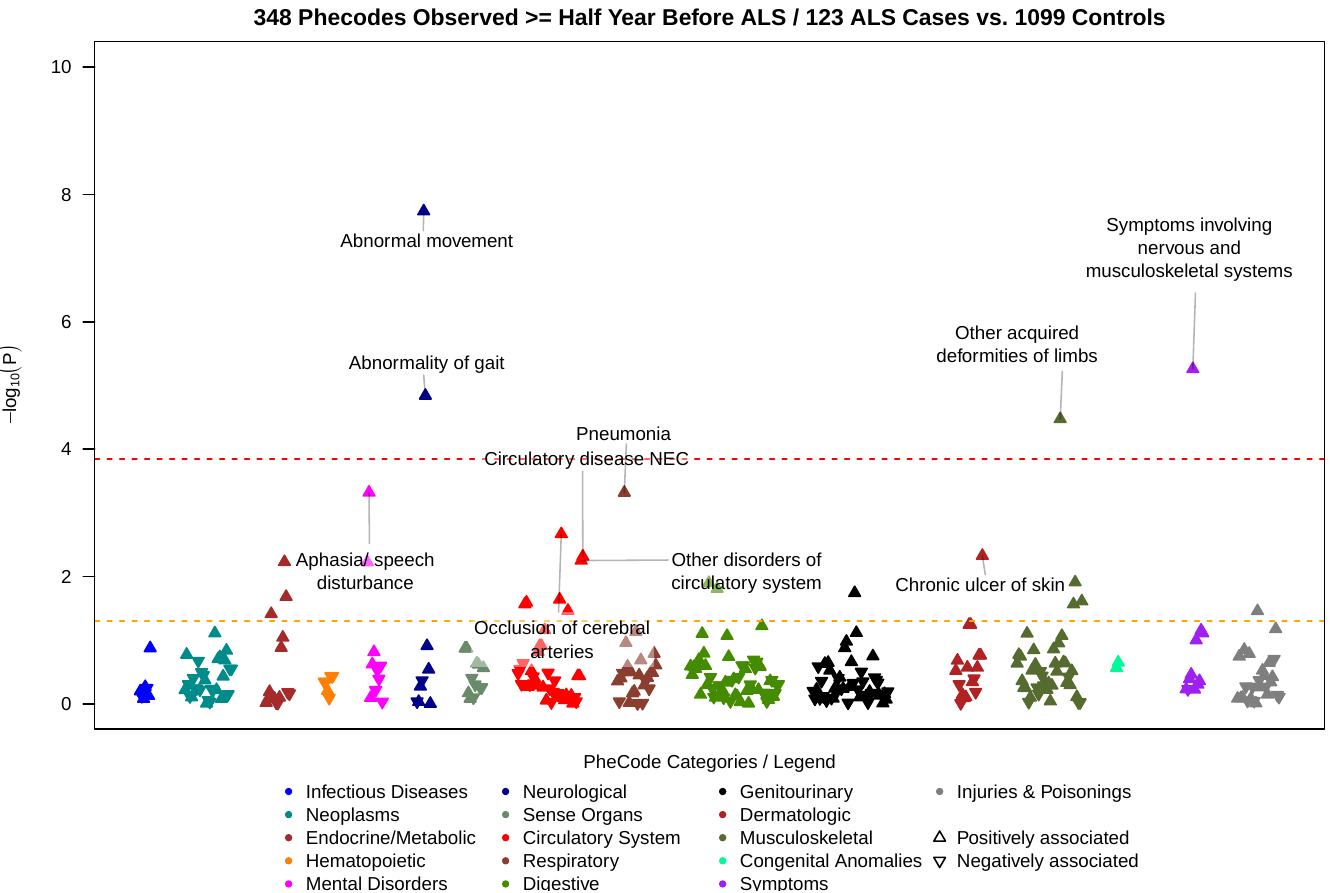

**B**

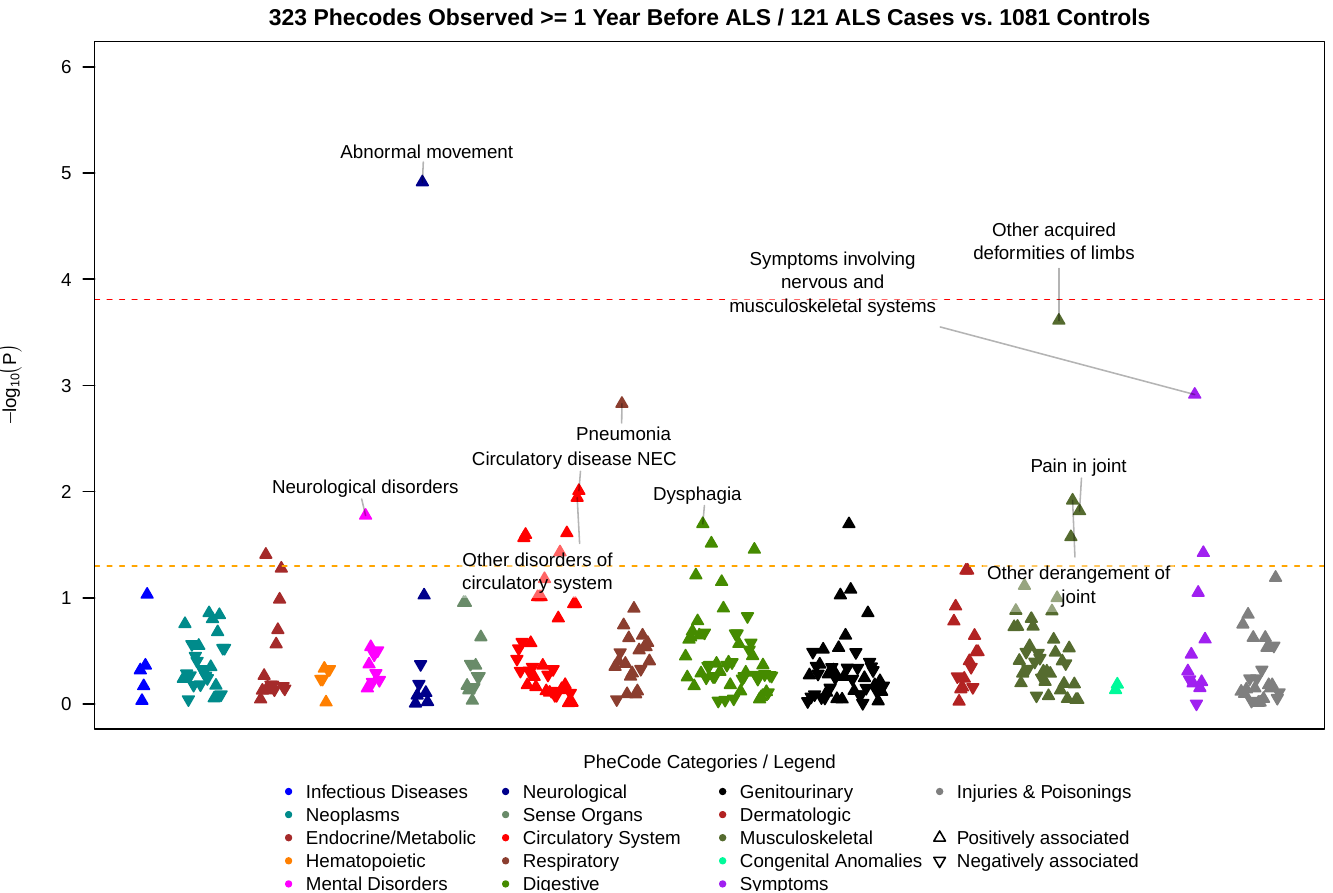

**C**

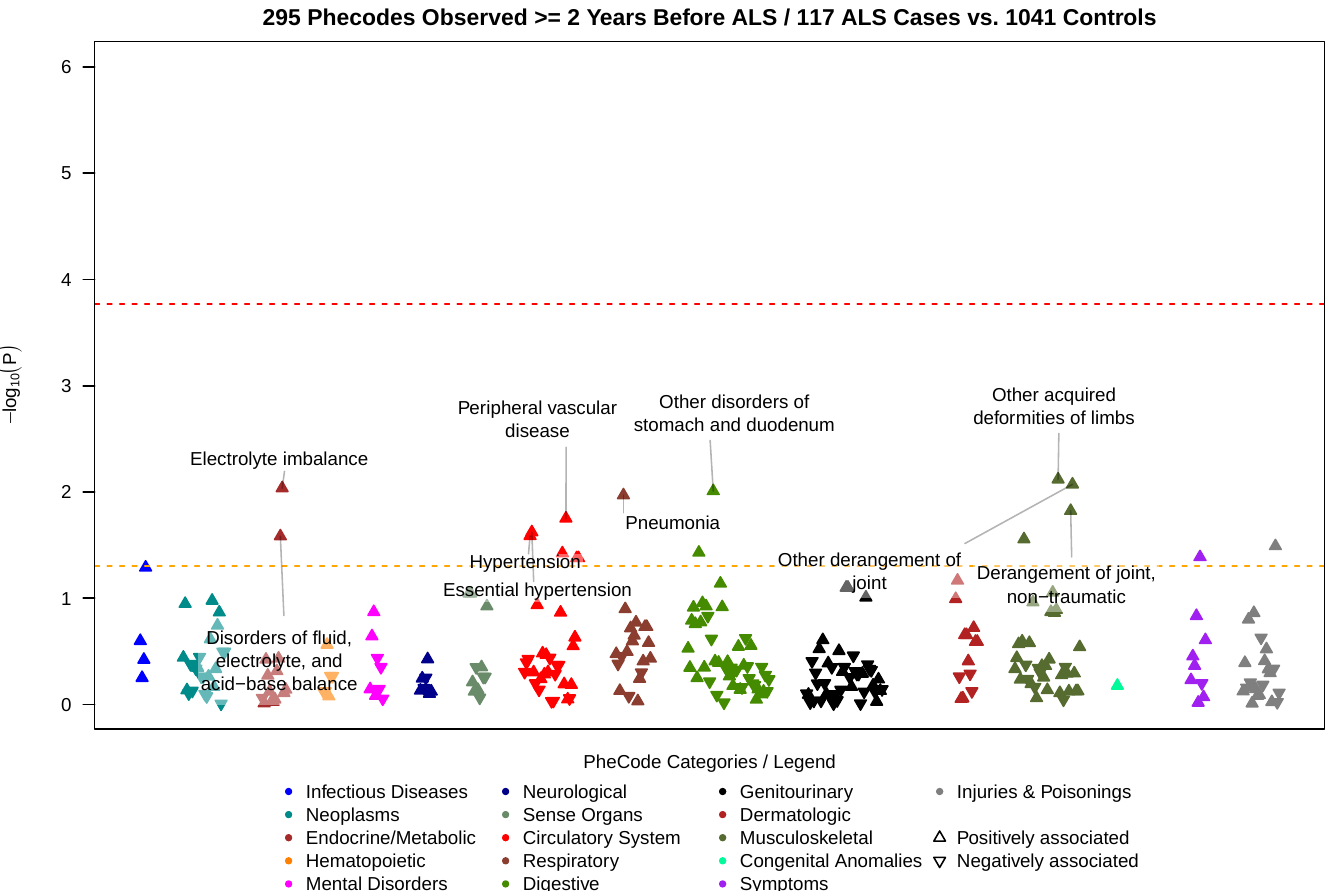

**D**

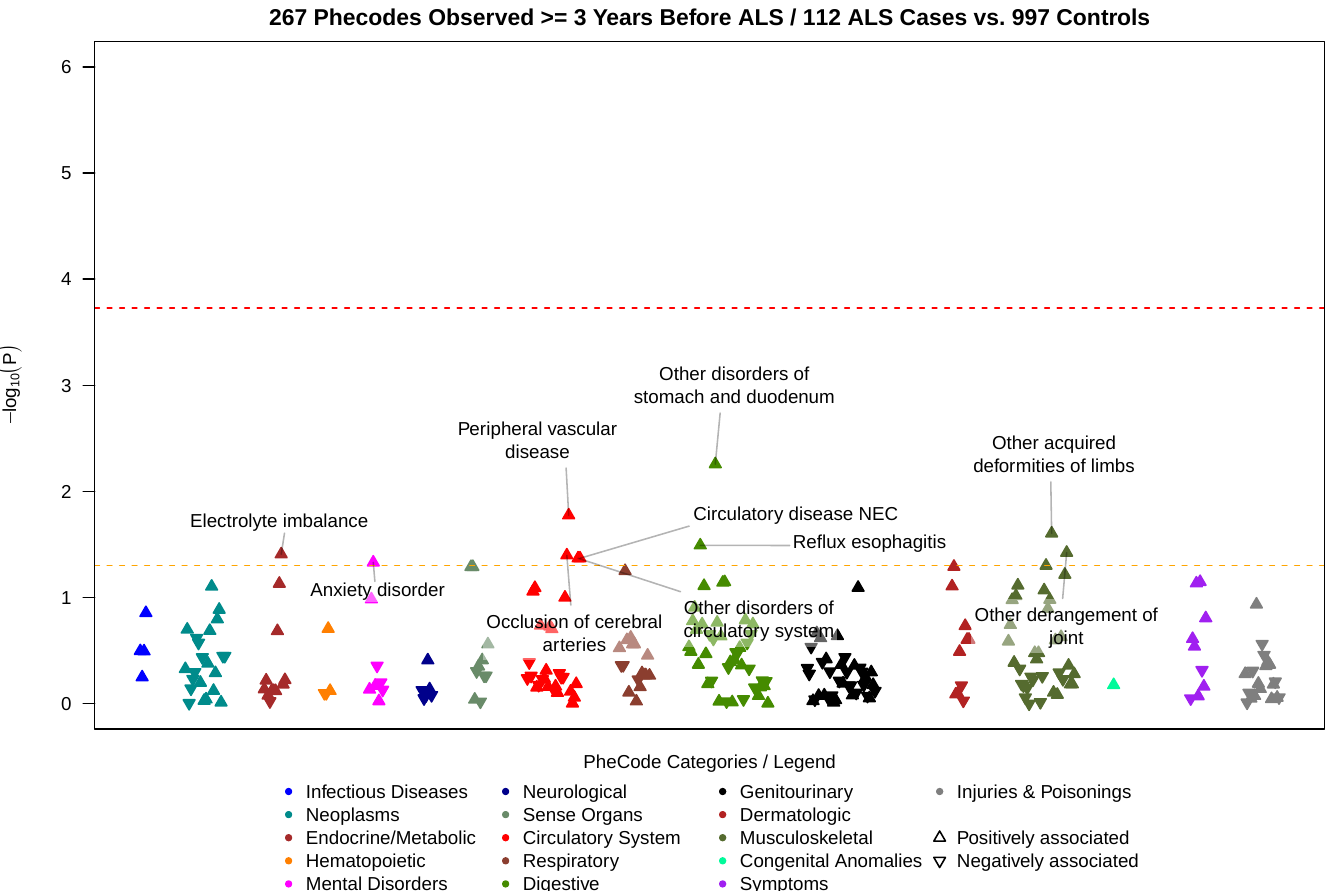

**E**

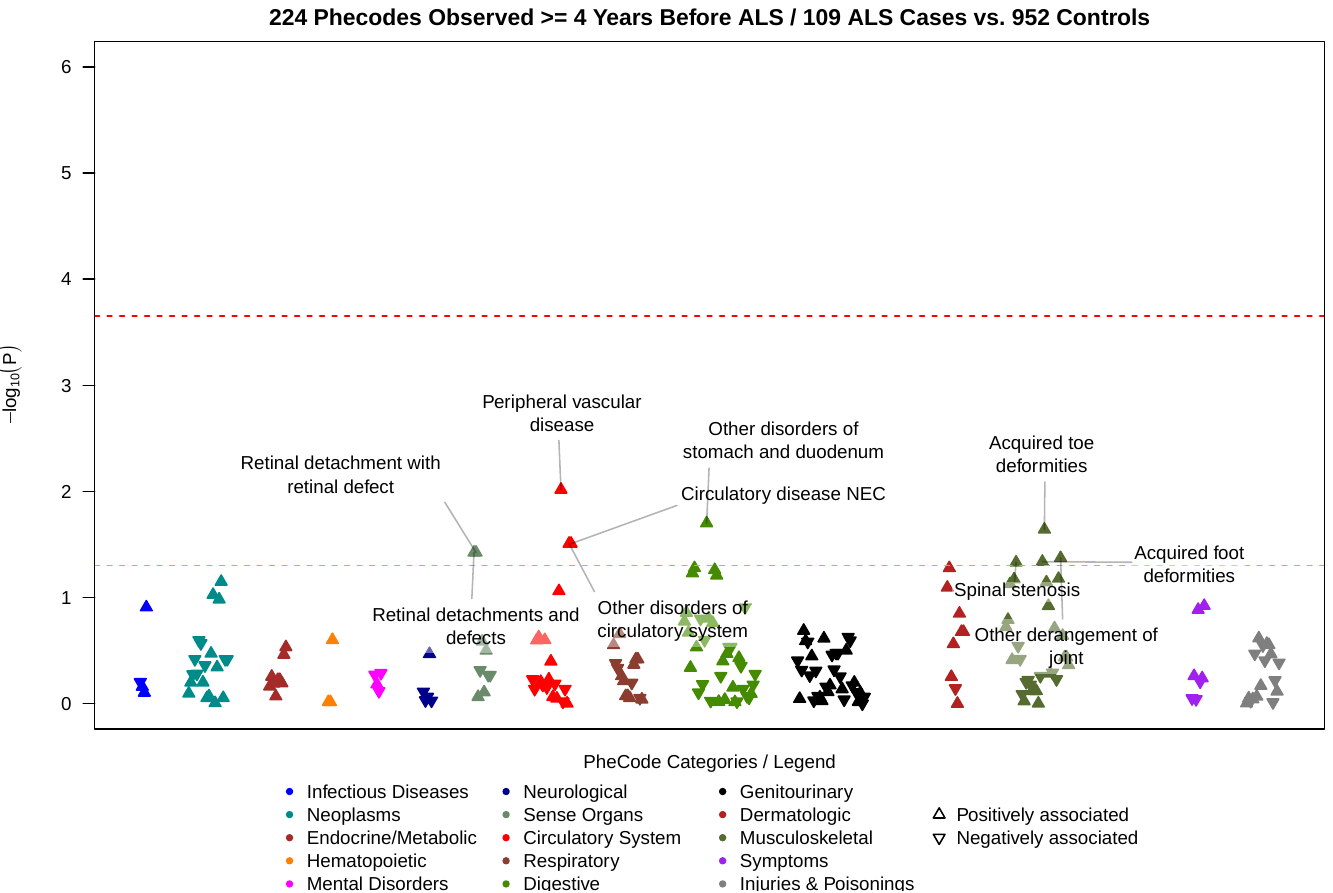

**F**

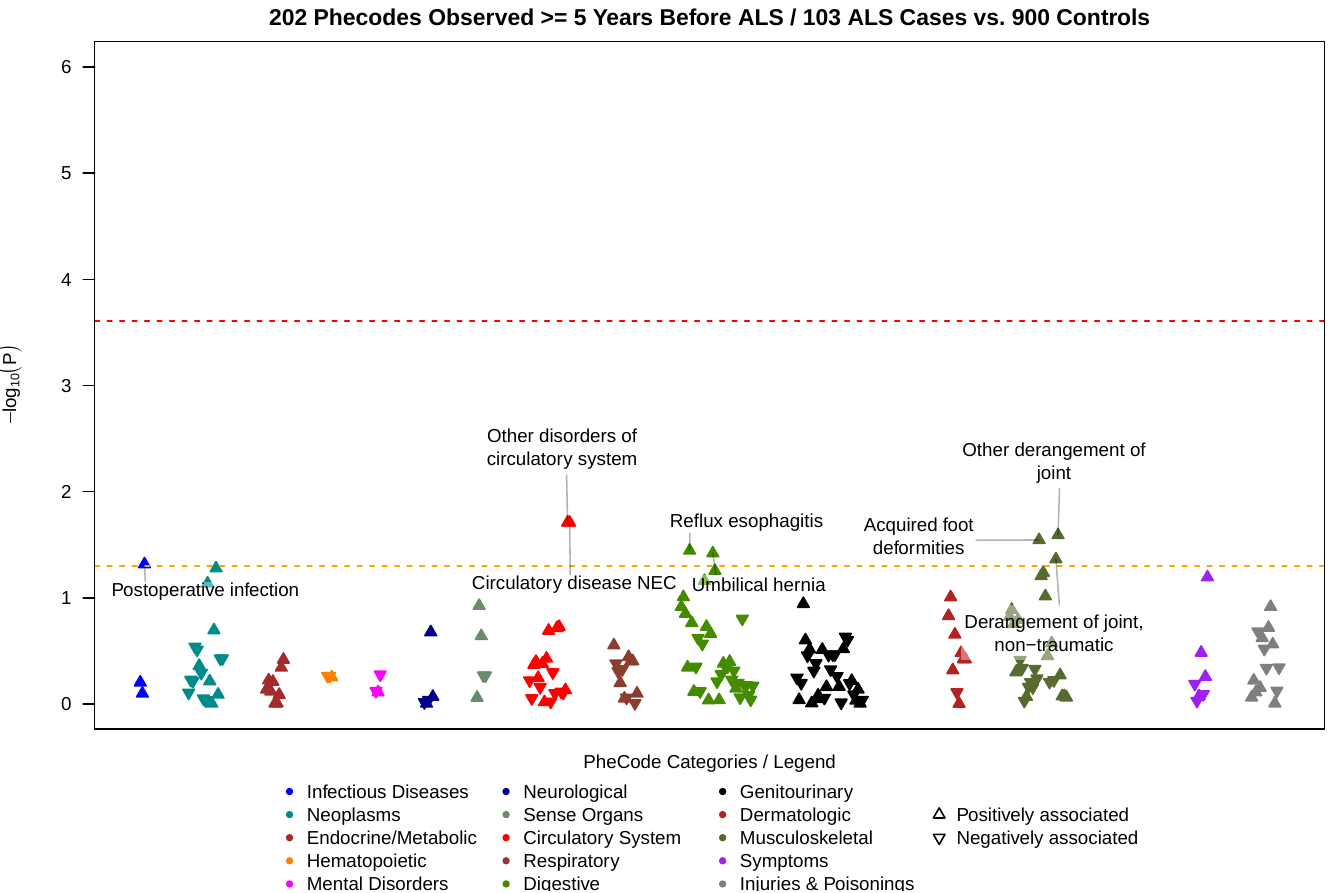
